# FACTORS AFFECTING PREMARITAL SICKLE CELL SCREENING UPTAKE AMONG PREGNANT WOMEN ATTENDING ANTENATAL CLINIC AT LIRA REGIONAL REFERRAL HOSPITAL: A CROSS-SECTIONAL STUDY

**DOI:** 10.1101/2025.08.03.25332899

**Authors:** Alice Naggujja, Jairus Wangusa Byawele, Maxson Kenneth Anyolitho, Emmanuel Ogwal, Akiror Bridget, Marc Sam Opollo

## Abstract

**Background:** Sickle cell disease (SCD), a genetic disorder marked by abnormal hemoglobin, poses a significant public health burden globally, affecting 20–25 million people, with a high prevalence and mortality in sub-Saharan Africa where over 300,000 infants are born annually with the disease. Uganda bears the fifth-highest global burden of SCD and ranks first in East Africa. Lira City reports a sickle cell trait prevalence of over 20% and a disease prevalence of 2%. The high teenage pregnancies in the region further underline the need for targeted premarital sickle cell screening.

**Objective:** To determine the level of uptake and factors affecting the premarital sickle cell screening among pregnant women aged 18 to 35 years attending antenatal clinic at Lira Regional Referral Hospital.

**Methods:** A hospital-based cross-sectional study was conducted among 170 pregnant women aged 18 to 35 years attending Antenatal care at Lira Regional Referral Hospital. Sample size was determined using the Kish-Leslie formula, 1965. Systematic random sampling was used to select study participants. Data were collected using a structured questionnaire administered via Kobo Collect and analysed using STATA version 18. Descriptive statistics, binary logistic regression were used at bivariate and multivariate level.

**Results:** Of the 170 respondents, only 14.7% had undergone premarital sickle cell screening. Uptake was significantly associated with having seen a PSCS-related poster (AOR = 7.71, 95% CI: 1.60–37.17) and having visited a health facility specifically for PSCS (AOR = 9.09, 95% CI: 15.54–54.22).

**Conclusion:** The uptake of Premarital Sickle Cell Screening (PSCS) was low at 14.7%, mainly due to limited awareness and poor health-seeking behavior. Exposure to PSCS posters and visits to health facilities were significantly associated with higher uptake. The study recommends increased sensitization and promotion of good health-seeking practices to improve screening rates.

## Background

Sickle cell disease (SCD) is a public health challenge globally affecting approximately 20-25 million people (1). Sickle cell disease is a genetic disorder characterized by abnormal haemoglobin in red blood cells. When two partners who both carry the sickle cell trait decide to have children, there’s a 25% chance that each pregnancy will result in a child with sickle cell disease. If one partner has sickle cell disease and the other is a carrier, the probability of having a child with sickle cell disease increases to 50% for each pregnancy (2).

In sub-Saharan Africa, more than 300,000 infants are born with sickle cell disease annually in West, Central, and Eastern Africa (3), In these Sub Saharan regions condition is often fatal in childhood, with an estimated under-5-year mortality rate ranging from 50% to 90%. Given the substantial impact SCD has on the community in the absence achieving Sustainable Development Goals (SDG) target 3.4 which aims at reducing one-third premature mortality from non-communicable diseases by 2030 and SDG target 3.2 that aims at end preventable deaths of newborns and children under 5 years of age by 2030 will be difficult to ascertain (4).

In Uganda, the overall uptake of PSCS is 22.8% and currently holds the fifth-highest global burden of sickle cell births and ranks first in the East African Subcontinent (5), with the prevalence of sickle cell trait in Uganda estimated at 13.3%, resulting in significant psycho-social and economic impacts on the patients, families and health sector at large (6). In the regions of Uganda the high prevalence of sickle cell disease contributes to a significant mortality rate, with approximately 25 percent of the 20,000 children born and diagnosed with sickle cell disease per year dying before reaching their first birthday (7). Despite the efforts by the Ministry of Health and partners such as establishing St. Mary’s Hospital Lacor as a Centre of Excellence, implementing pilot neonatal haemoglobinopathy screening, providing hydroxyurea treatment, and conducting awareness campaigns, these interventions have mainly improved disease management and early detection. However, they have not significantly impacted prevention, as the uptake of premarital sickle cell screening remains low at 22% and the prevalence of sickle cell trait in Uganda remains high at 13.3%.(8).

A study done in Hoima, Uganda on the utilization of preconception sickle cell screening reveals that counseling and hereditary screening services are available in numerous facilities and institutions across various cities and countries. However, the availability of preconception and premarital screening and counseling for genetic disorders is limited. Preconception counseling and testing are considered strategic measures to reduce the burden of sickle cell disease, as evidenced in high-endemicity areas like Cyprus and Sardinia (9).

Mid-northern Uganda where Lira city is situated presents significant burdens of both sickle cell trait (>18%) and sickle cell disease (>1%) (10). The prevalence of SCD in Lira City remains high at 2% and SCT at >20%(11) According to recent reports from LRRH , approximately 2 out of 100 children born in the Lira Regional Referral hospital are affected by sickle cell disease (12). Previous studies conducted in and outside Uganda about sickle cell disease have found out that factors associated with uptake of sickle cell screening include awareness, age, the availability and cost of screening services, individual perceptions of SCD, traditional beliefs, and family history of sickle cell disease (13) (14) (15) (16).

According to a study conducted by Ochen et al. (2019) in Lira City, Northern Uganda, 90.3% of girls become pregnant between the ages of 13 and 19. These findings highlight the urgent need to strengthen screening programs among adolescents to identify carriers and reduce the transmission of sickle cell disease. Enhancing healthcare services, particularly premarital screening, alongside community awareness initiatives, is essential for managing and preventing complications. Such efforts can help shift attitudes and behaviors, enabling young couples to make informed reproductive choices and secure better health outcomes for future generations. (17).

This study was set to investigate factors affecting PSCS uptake among pregnant women attending Antenatal care services at Lira Regional referral hospital, generating new knowledge to inform evidence-based recommendations for policies, including advocating for mandatory premarital screening to enhance uptake and reduce the burden of SCD.

## Methods and Materials

### Study Design

A cross-sectional study design employing a quantitative data collection method was used, and data were collected from 11^th^ February to 11^th^ May 2025. This design allowed for the collection of data at a single point in time, providing a snapshot of the uptake and related factors of premarital sickle cell screening.

### Study Setting

The study was conducted at Lira Regional Referral Hospital (LRRH), located in Lira City West Division, approximately 340 kilometres north of Kampala. LRRH is a major referral hospital in the Lango Sub-region, serving nine districts and one city, with an annual patient load of about 2.5 million. The hospital provides both specialized and general healthcare services and offers antenatal care to approximately 80 women daily. The study took place in the maternal and child health clinic within the Obstetrics and Gynaecology department.

### Study Participants

The target population included all prime gravida women. The study focused on married prime gravida aged 18 to 35 years who were attending antenatal care services at LRRH.

Women were eligible for inclusion if they were married, aged between 18 and 35 years, and attending ANC during the study period. Those who were too ill to provide information were excluded.

### Sample Size

The sample size was calculated using the Kish-Leslie formula (1965) for cross-sectional studies. With a confidence interval of 95% (Z = 1.96), an estimated prevalence of premarital sickle cell screening at 11.4% (p = 0.114), a complementary proportion (q = 1 − p = 0.886), and a margin of error (e) of 5%, the computed sample size was 155. To account for a 10% non-response rate, 16 more participants were added, yielding a final sample size of 171.

### Sampling Procedure

Systematic random sampling was used to select participants. The sampling frame included all eligible women attending ANC during the study period. To select the first participant, a lottery method was employed using the first three names in the registration system (EPHIA). Following this, every third eligible woman was selected based on the calculated sampling interval (560 ÷ 171 = 3).

### Measurements of Study Variables

The outcome variable was premarital sickle cell screening, defined as whether a participant had undergone sickle cell screening before marriage (coded as 1 = Yes, 0 = No). Independent variables were grouped into sociodemographic factors, individual knowledge and perceptions, and health system–related factors. Examples included age, education, occupation, awareness of PSCS and SCD, beliefs about SCD, previous exposure to health information, and health facility visits related to screening.

### Data Collection Process

Data were collected using a pretested structured, interviewer-administered questionnaire developed in English and translated into Leb Lango to enhance comprehension. The questionnaire had four sections: socio-demographic characteristics, uptake of PSCS, individual-level factors, and health system–related factors. Interviews were conducted privately in a comfortable space, lasting between 12 and 42 minutes.

The Kobo Collect mobile application was used to collect and upload data in real time. Research assistants fluent in Leb Lango and trained in research ethics and tool administration facilitated the process. Participants were interviewed only after obtaining verbal and written informed consent. A transport refund was provided after completion of the interview. The entire data collection process lasted six days.

### Data Analysis and Statistical Methods

Data collected via Kobo Collect were exported to Microsoft Excel, cleaned, coded, and analyzed using STATA version 18. Descriptive statistics were used to summarize participants’ characteristics and levels of PSCS uptake using frequencies and percentages. Factors associated with PSCS uptake were assessed using bivariate and multivariate logistic regression. Variables with the P value < 0.2 at bivariate level were included in the multivariable model. Confounding and interaction were evaluated and variables with a p value < 0.05 were considered statistically significant.

## RESULTS

A total of 220 married prime gravida women attending ANC at Lira Regional Referral Hospital (LRRH) were screened for eligibility. Among these, 50 were excluded based on the exclusion criteria. The final sample comprised 170 participants, resulting in a response rate of 99.4%.

### Uptake of Premarital Sickle Cell Screening (PSCS)

Out of the 170 study participants, only 25 (14.7%) reported having undergone premarital sickle cell screening prior to marriage. This is illustrated in Figure 2.

**Figure 1:**
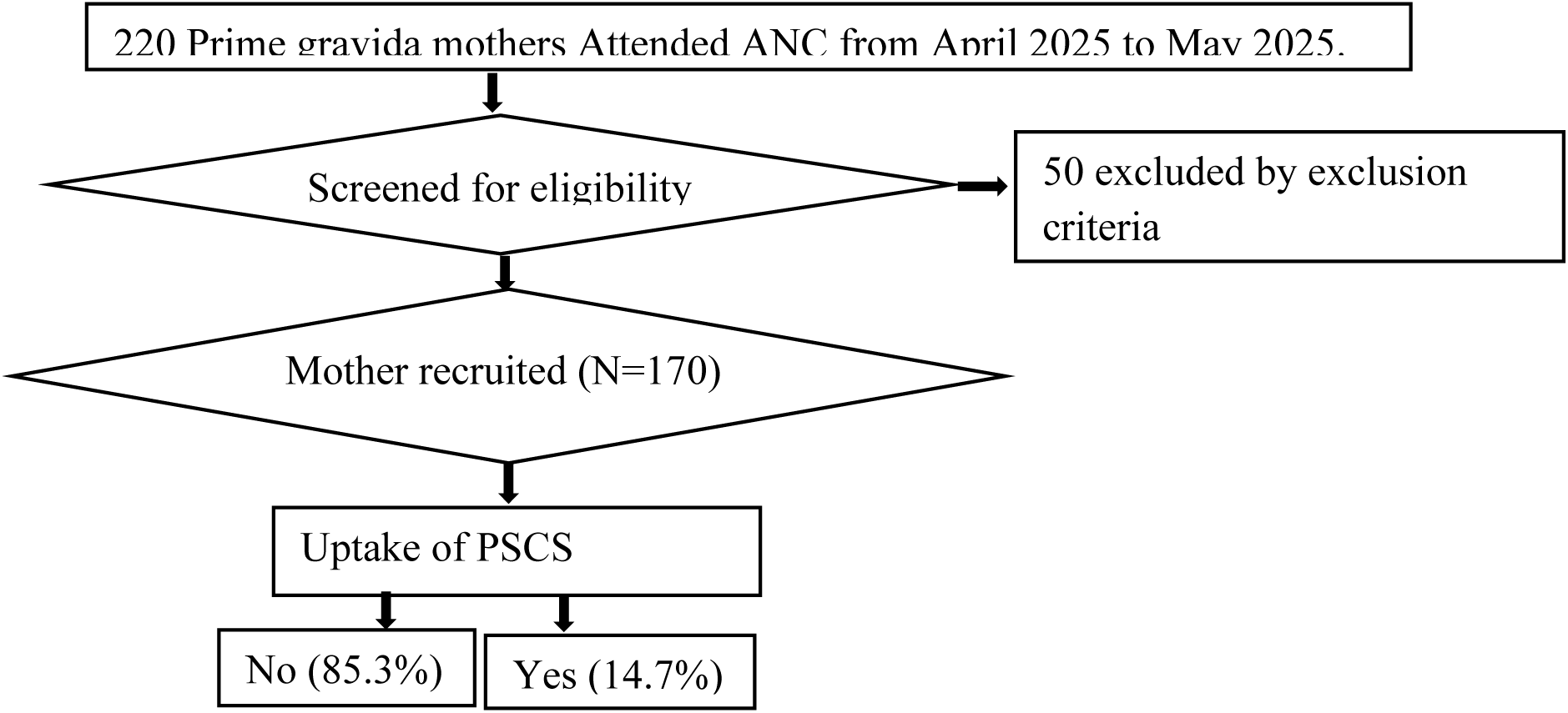
Study profile of pregnant women attending ANC at LRRH

**Figure 2:**
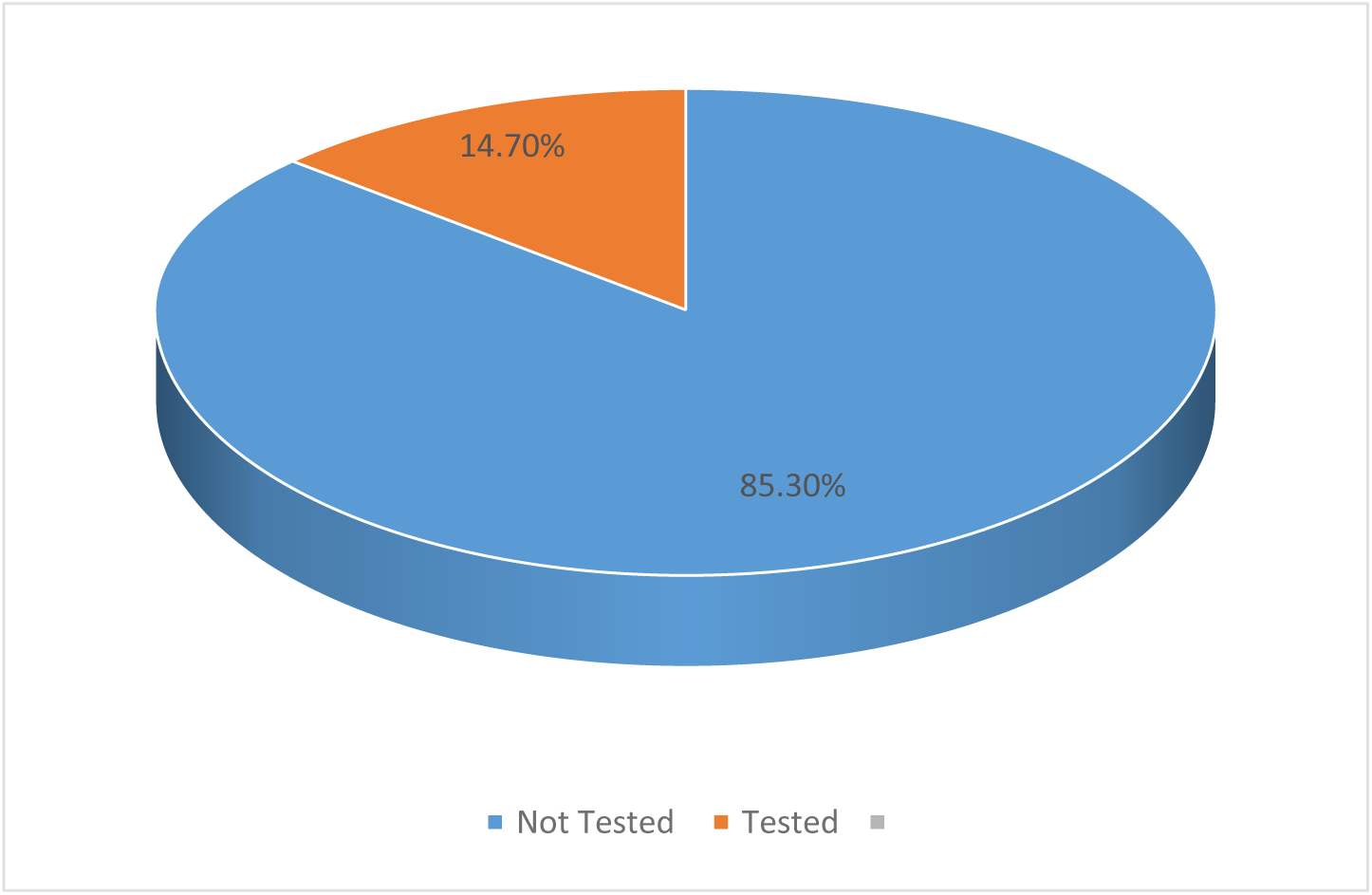
Uptake of PSCS among pregnant women attending ANC at LRRH.

### Socio-demographic Characteristics

The majority of participants (92; 54.1%) were aged between 24 and 29 years. Most identified as Catholic (81; 47.6%), had attained secondary education (90; 52.9%), and were self-employed (70; 41.2%). These details are summarized in Table 1.

**Table 1:** Socio-demographic Characteristics of Study Participants (n=170).

| Variable | Frequency | Percentage (%) |
| --- | --- | --- |
| **Age** |  |  |
| 18-23 | 2 | 30.59 |
| 24-29 | 92 | 54.12 |
| 30-35 | 26 | 15.29 |
| **Religion** |  |  |
| Catholic | 81 | 47.65 |
| Protestant | 55 | 32.35 |
| Moslem | 20 | 11.76 |
| Born Again | 13 | 7.65 |
| SDA | 1 | 0.59 |
| **Highest Education Level** |  |  |
| Primary | 45 | 26.47 |
| Secondary | 90 | 52.94 |
| Tertiary/University | 35 | 20.59 |
| **Current Occupation** |  |  |
| Self-employed | 70 | 41.18 |
| Casual laborer | 28 | 16.47 |
| Student | 25 | 14.71 |
| Formal employment | 24 | 14.12 |
| Unemployed | 23 | 13.53 |

### Knowledge and Awareness of PSCS and SCD

Out of 170 participants, 111 (65.3%) had ever heard of PSCS, while 164 (96.5%) had heard of sickle cell disease (SCD). Among those who had heard of PSCS, the main sources of information were healthcare providers (47; 42.3%) and schools (45; 40.5%). A majority (154; 90.6%) rejected the misconception that SCD affects only females, and 164 (96.5%) believed PSCS results should be considered in marital decision-making. Only 37 (21.8%) correctly identified that SCD can be asymptomatic. These findings are presented in Table 2.

**Table 2:** Knowledge and Awareness of PSCS (n=170).

| **Variables** | **Frequency** | **Percentage (%)** |
| --- | --- | --- |
| **Ever Heard of PSCS** |  |  |
| Yes | 111 | 65.29 |
| No | 58 | 34.12 |
| **Main Source of PSCS Info** |  |  |
| Healthcare provider | 47 | 42.34 |
| School | 45 | 40.54 |
| TV/Radio | 9 | 8.11 |
| Friends/Family | 9 | 8.11 |
| Don't remember | 1 | 0.90 |
| **Ever Heard of SCD** |  |  |
| Yes | 164 | 96.47 |
| No | 6 | 3.53 |
| **Belief: SCD Affects Only Females** |  |  |
| Wrong | 154 | 90.59 |
| Right | 16 | 9.41 |
| **Belief: SCD Can Be Asymptomatic** |  |  |
| Right | 37 | 21.76 |
| Wrong | 133 | 78.24 |
| **Belief: PSCS Result Used for Marriage Decision** |  |  |
| Right | 164 | 96.47 |
| Wrong | 6 | 3.53 |

### Bivariate Analysis – Individual-Level Factors

Bivariate logistic regression identified education level, awareness of PSCS, and knowledge that SCD can occur with symptoms as significant factors (p < 0.2). Participants with tertiary/university education were 5.6 times more likely to have undergone PSCS compared to those with primary education (COR = 5.600, 95% CI: 1.430–21.934, p = 0.013). Respondents who had ever heard of PSCS were 5.8 times more likely to have been screened (COR = 5.848, 95% CI: 1.310–26.117, p = 0.021). Awareness that SCD can occur with symptoms was associated with a 2.3-fold higher likelihood of PSCS uptake (COR = 2.349, 95% CI: 1.128–4.894, p = 0.023). These associations are summarized in Table 3.

**Table 3:** Bivariate Analysis of Association between Individual Factors and PSCS (n=170)

| Variable | Tested PSCS |  | COR (95% CI) | p-value |
| --- | --- | --- | --- | --- |
|  | No | Yes |  |  |
| Age group |  |  |  |  |
| 18–23 | 45 | 4 | 1.00 |  |
| 24–29 | 78 | 15 | 1.313 (0.487–3.538) | 0.590 |
| 30–35 | 22 | 6 | 1.540 (0.455–5.211) | 0.492 |
| Education level |  |  |  |  |
| Primary | 52 | 4 | 1.00 |  |
| Secondary | 70 | 12 | 2.229 (0.683–7.270) | 0.286 |
| Tertiary/University | 23 | 9 | 5.600 (1.430–21.934) | 0.013* |
| **Occupation** |  |  |  |  |
| Government-employed | 17 | 5 | 1.00 |  |
| Private-employed | 25 | 7 | 0.952 (0.215–4.206) | 0.949 |
| Self-employed | 47 | 8 | 1.240 (0.379–4.055) | 0.721 |
| Unemployed | 56 | 5 | 0.905 (0.186–4.394) | 0.902 |
| **Heard of PSCS** |  |  |  |  |
| No | 44 | 3 | 1.00 |  |
| Yes | 101 | 22 | 5.848 (1.310–26.117) | 0.021* |
| **Knows SCD can occur with symptoms** |  |  |  |  |
| No | 77 | 10 | 1.00 |  |
| Yes | 68 | 15 | 2.349 (1.128–4.894) | 0.023* |
| **Believes PSCS result should influence marriage** |  |  |  |  |
| No | 45 | 9 | 1.00 |  |
| Yes | 100 | 16 | 0.800 (0.363–1.762) | 0.578 |
| **Interested in learning more about PSCS** |  |  |  |  |
| No | 36 | 5 | 1.00 |  |
| Yes | 109 | 20 | 1.322 (0.514–3.399) | 0.561 |

### Bivariate Analysis of Health System Factors

Bivariate analysis of health system variables showed that visiting a health facility for PSCS (COR = 3.11, 95% CI: 1.020–2.777, p = 0.07) and seeing a poster about PSCS (COR = 2.534, 95% CI: 1.755–3.117, p = 0.05) were associated with higher screening uptake. These are presented in Table 4.

**Table 4:**
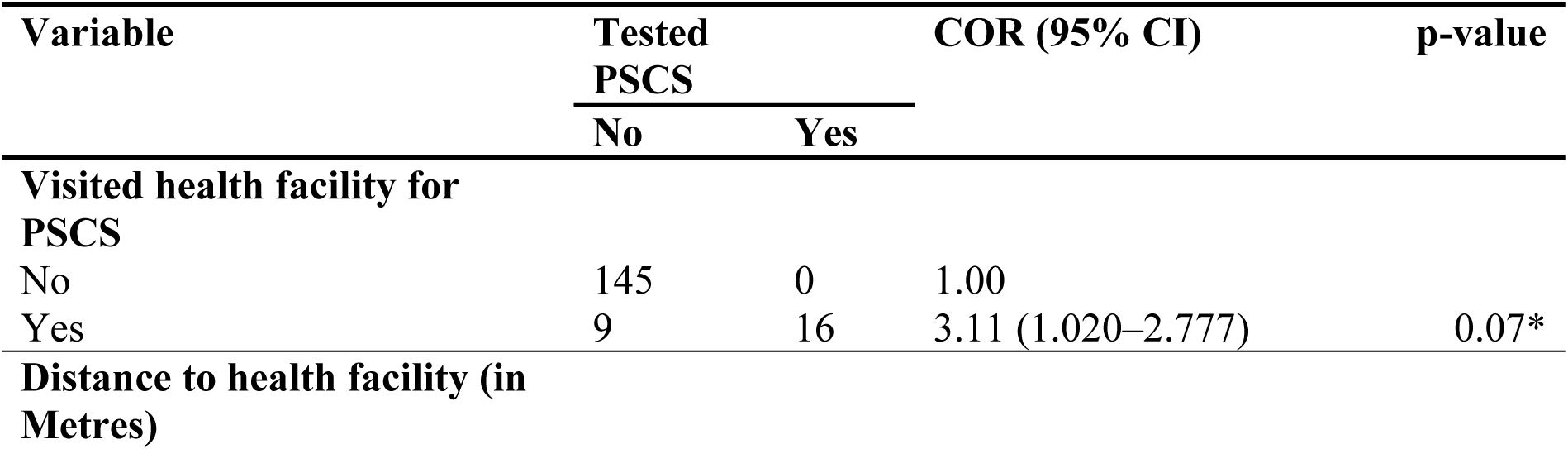
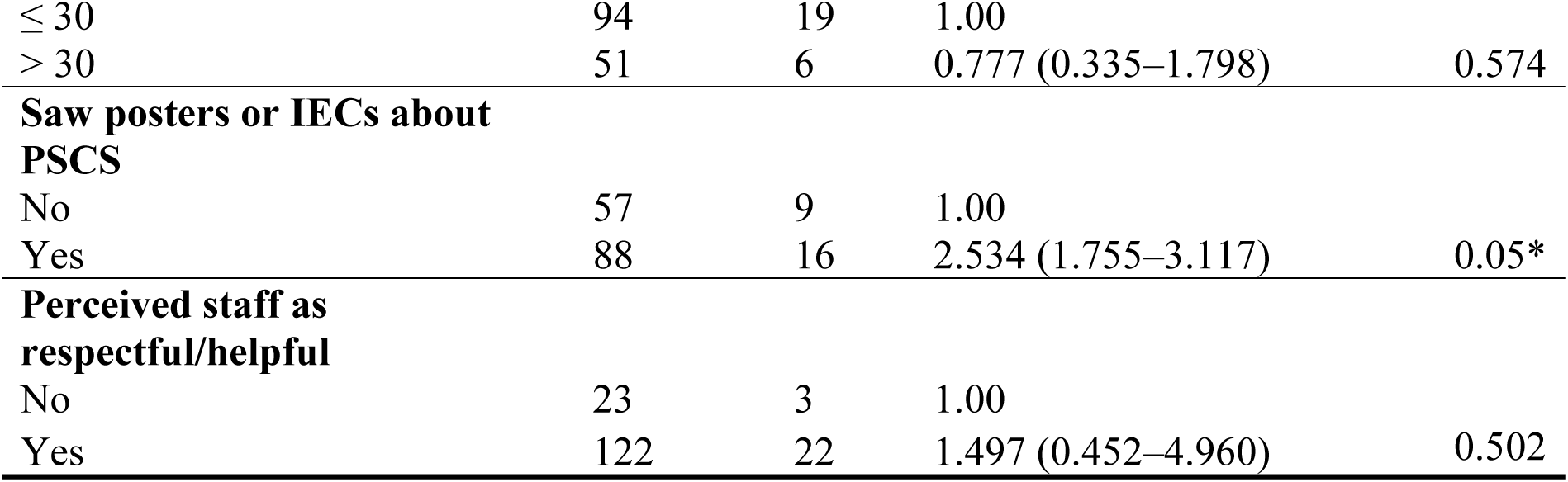
Bivariate Analysis of Health System Factors Associated with PSCS (n=170)

### Multivariate Analysis of Factors Affecting the Premarital Sickle Cell Screening Uptake

Multivariate logistic regression was conducted to determine the independent predictors of PSCS uptake. Variables that had p < 0.2 at the bivariate level were included in the model. The final model revealed that respondents who had seen a poster about PSCS were 7.7 times likely to have undergone screening than those who did not see the posters (AOR = 7.71, 95% CI: 1.60–37.17, p = 0.011), and those who had visited a health facility specifically for PSCS were 9.1 times more likely to have been screened (AOR = 9.09, 95% CI: 5.54–59.22, p < 0.001) these were presented in table 5.

**Table 5:**
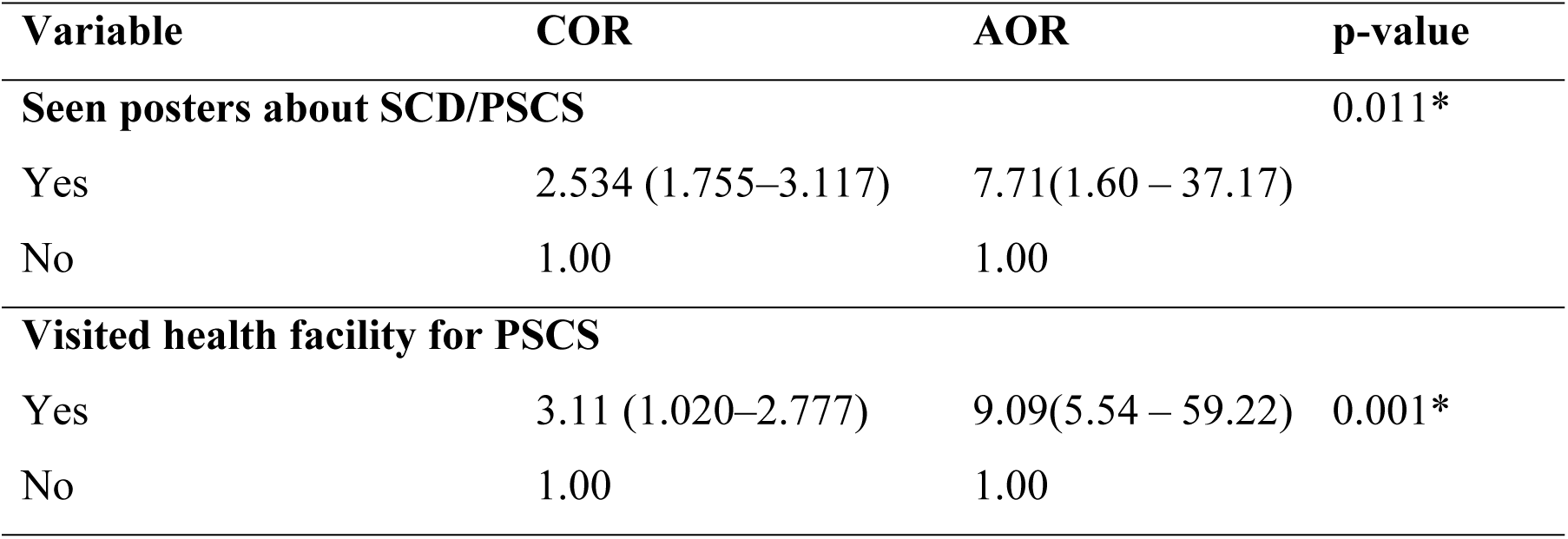
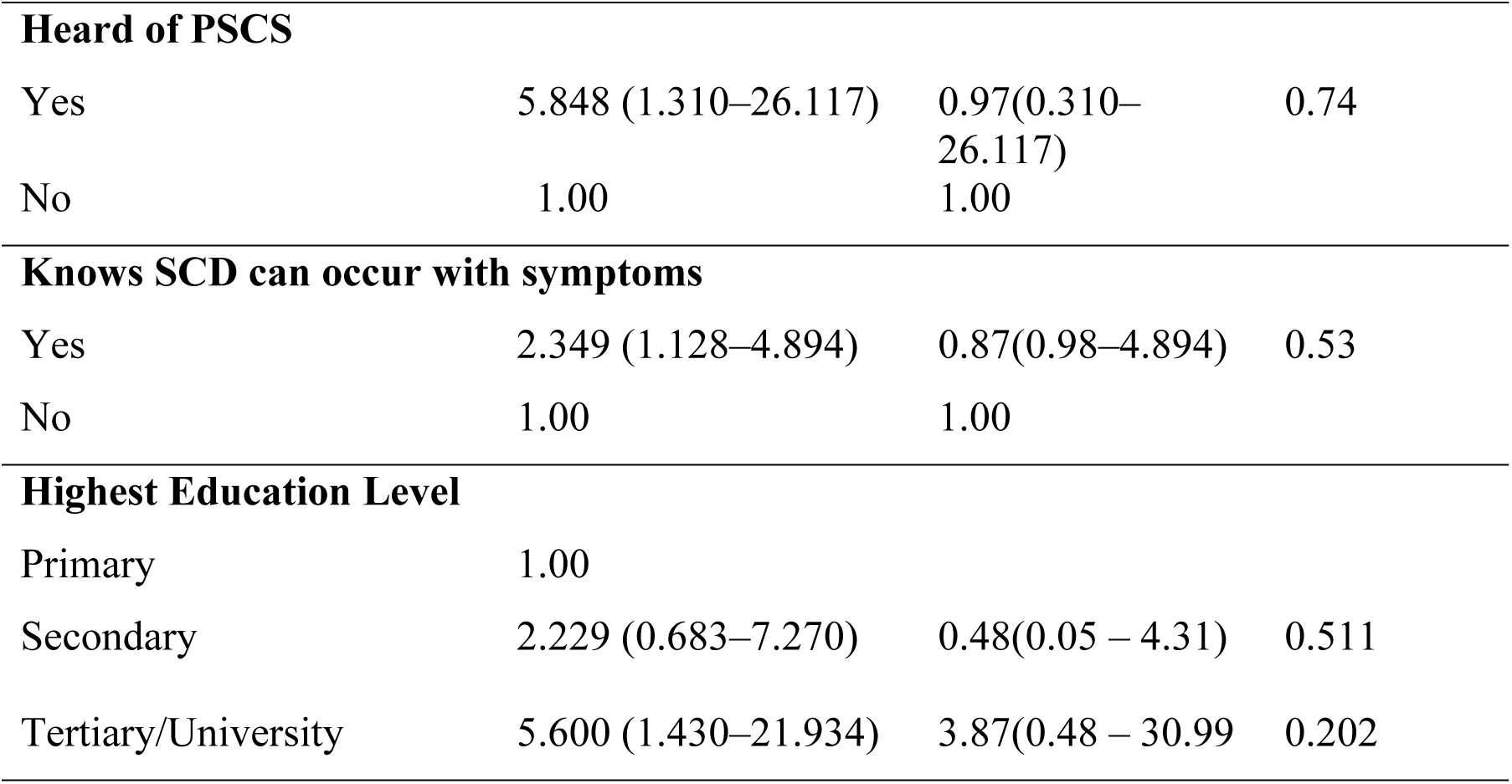
Multivariate Analysis of Factors Associated with PSCS Uptake among Married Prime Gravida (n=170).

| Variable | COR | AOR | p-value |
| --- | --- | --- | --- |
| **Seen posters about SCD/PSCS** |  |  | 0.011* |
| Yes | 2.534 (1.755–3.117) | 7.71(1.60 – 37.17) |  |
| No | 1.00 | 1.00 |  |
| **Visited health facility for PSCS** |  |  |  |
| Yes | 3.11 (1.020–2.777) | 9.09(5.54 – 59.22) | 0.001* |
| No | 1.00 | 1.00 |  |
| **Heard of PSCS** |  |  |  |
| Yes | 5.848 (1.310–26.117) | 0.97(0.310–26.117) | 0.74 |
| No | 1.00 | 1.00 |  |
| **Knows SCD can occur with symptoms** |  |  |  |
| Yes | 2.349 (1.128–4.894) | 0.87(0.98–4.894) | 0.53 |
| No | 1.00 | 1.00 |  |
| **Highest Education Level** |  |  |  |
| Primary | 1.00 |  |  |
| Secondary | 2.229 (0.683–7.270) | 0.48(0.05 – 4.31) | 0.511 |
| Tertiary/University | 5.600 (1.430–21.934) | 3.87(0.48 – 30.99) | 0.202 |

## Discussions of Key Findings

This study found a low uptake of premarital sickle cell screening (PSCS), with only 14.7% of married prime gravida women aged 18–35 at Lira Regional Referral Hospital (LRRH) having undergone PSCS prior to marriage. This result highlights a critical gap in the implementation of preventive strategies for sickle cell disease (SCD) in Uganda. The low uptake may be attributed to the voluntary nature of PSCS in Uganda, insufficient health education during adolescence and early adulthood, and limited integration of screening into premarital or antenatal care services. The consequence of such low uptake is continued risk of SCD transmission among couples unaware of their genotype status, contributing to the national burden of the disease. This finding aligns with a national study by Olupot-Olupot et al. (2020), which reported a PSCS uptake of 22%, and is comparable to Ochen et al. (2019), who noted limited premarital screening among adolescents in Lira. In contrast, countries like Nigeria report uptake as high as 86.7% due to religious and legal mandates requiring proof of screening before marriage (18). This underscores how mandatory policies can significantly improve uptake.

In this study, education level significantly influenced PSCS uptake. Women with tertiary education were 5.6 times more likely to have undergone PSCS compared to those with only primary education (COR = 5.600, p = 0.013). This finding reflects the role of education in improving health literacy, awareness of genetic conditions, and ability to make informed health decisions. Educated women may also have better access to health information through schools and media, and may be more empowered to negotiate screening with their partners. The implication is that interventions promoting PSCS should incorporate educational outreach, especially targeting low-literate populations. This is consistent with findings by Dilli et al. (2024) and Olupot-Olupot et al. (2020), who both noted higher PSCS uptake among those with secondary or tertiary education in Uganda. Similarly, studies from Nigeria and Ghana have shown that educated women are more likely to understand the implications of SCD and seek premarital testing (13) (18).

The study also revealed that women who had heard of PSCS were 5.8 times more likely to be screened than those who had not (COR = 5.848, p = 0.021), and those who knew that SCD can occur with symptoms were 2.3 times more likely to be screened (COR = 2.349, p = 0.023). This suggests that awareness and accurate knowledge of SCD and PSCS are key enablers of screening behavior. Lack of awareness contributes to misconceptions and a diminished perceived need for testing, reducing the likelihood of uptake. The impact of such gaps in knowledge is the persistence of preventable cases of SCD due to uninformed marital decisions. These findings echo those of Dilli et al. (2024), who found that awareness of SCD significantly predicted screening behavior. Similar associations have been observed in school-based studies in Uganda (19) (6), where students with better knowledge were significantly more likely to test.

Although not statistically significant, the results showed that younger women (18–23 years) were less likely to have undergone PSCS compared to older age groups. Younger women may have less exposure to reproductive health education, limited autonomy in making health decisions, or may enter marriage early without adequate premarital counseling. This pattern suggests that adolescent-friendly health services and youth-centered awareness campaigns are needed to reach this high-risk group. These findings are consistent with Agbozo et al. (2023), who found that women under 20 were four times less likely to test than those over 30. Kisakye et al. (2022) similarly reported low PSCS uptake among students under 25, despite moderate levels of knowledge.

Occupation was not significantly associated with PSCS uptake in this study, but descriptively, self-employed and government-employed women had higher screening rates than casual laborers or unemployed women. This could be due to better financial stability, access to health services, and greater exposure to health information among the employed. Conversely, unemployed or casually employed women may face financial constraints that prevent them from accessing screening services. These findings are consistent with Olupot-Olupot et al. (2020), who found that financial barriers deter screening uptake in Uganda. Similar trends have been reported in Nigeria and Ghana, where women in formal employment were more likely to be screened due to affordability and institutional health programs (20) (13).

The study revealed that visiting a health facility specifically for PSCS significantly increased the likelihood of screening (AOR = 9.09, p < 0.001). This suggests that active engagement with health services is a strong enabler of uptake. The likely reason is that visits to a facility expose women to counseling and diagnostic services, increasing awareness and motivation to test. The implication is that integrating PSCS services into routine care and improving their availability at lower-level facilities can boost uptake. This finding is in line with the conclusions of Gosadi et al. (2021) and Kyakuwa et al. (2024b), who identified the availability and integration of services as a key determinant of utilization. In Uganda, Dilli et al. (2024) similarly found that many women who tested had done so during clinic visits, where services were accessible.

Respondents who had seen posters or IEC materials related to PSCS were 7.7 times more likely to have tested (AOR = 7.71, p = 0.011). This result underscores the importance of health communication in influencing behavior. Posters serve as visual reinforcements that increase awareness, prompt action, and normalize screening. The impact of this finding is the justification for strengthening health education through visual aids in health facilities and communities. This aligns with studies by Anderson et al. (2021) and Mwangi et al. (2022), which found that educational posters enhance understanding and drive health-seeking behavior. In Lira, Kyakuwa et al. (2024b) also observed that exposure to PSCS messages through visual aids significantly increased uptake among pregnant women.

Distance to the nearest health facility was not significantly associated with PSCS uptake. However, women living more than 3 km away were less likely to have tested, suggesting that physical access may still pose a barrier. The implication is that outreach and mobile screening services could address access disparities, particularly for rural populations. These results are comparable to studies from Nigeria and Tanzania, which highlight distance and transport costs as key barriers to screening (21) (11). In Uganda, Baker et al. (2015) also noted that women in remote areas face reduced access to essential preventive services, including SCD screening.

Although most respondents (81.2%) reported receiving good treatment from healthcare providers, this factor was not significantly associated with PSCS uptake. Nevertheless, positive provider attitudes may influence other health behaviors, including willingness to return to the facility or recommend services to others. Provider engagement particularly through direct discussion and recommendation has been shown to increase screening adherence (8). In Uganda, Kyakuwa et al. (2024b) found that health worker counseling significantly predicted the intention to test for SCD among antenatal clients. The lack of significance in this study may be due to variability in the quality or depth of discussions held.

## Limitations of the Study

Cross-Sectional Study Design while the cross-sectional design was logistically feasible, it inherently limits the ability to infer causality or temporal relationships. Therefore, although associations were found between some health system factors (e.g., seen a poster and visiting the health facility) and uptake of PSCS, cannot be conclusively determined whether this uptake directly caused by these factors. However, this was mitigated by using multivariate logistic regression to control for confounders and by recruiting a diverse sample from a major referral hospital serving both urban and rural populations. Data on socioeconomic factors and health system experiences were obtained through self-report from study participants, this was minimized by using neutral, non-leading questions and ensuring privacy during data collection.

## Conclusion

This study revealed a low uptake, underscoring a critical gap in the implementation of preventive strategies for sickle cell disease (SCD) in Uganda. Factors such as tertiary education, prior awareness of PSCS, knowledge that SCD can be asymptomatic, visiting a health facility specifically for PSCS and exposure to educational posters showed the strongest associations with screening uptake. . These findings emphasize the role of health literacy, personal understanding in shaping health-seeking behavior, influence of accessible and engaging health services and communication strategies in promoting preventive action.

To improve uptake of Premarital Sickle Cell Screening (PSCS), it should be integrated into antenatal care and youth-friendly services. Health education and awareness campaigns must be intensified using visual materials and community engagement. Training health workers and equipping facilities will ensure consistent service delivery. Decentralizing services and removing user fees can address access and affordability barriers. National policy development and further research are essential for sustainable, evidence-based implementation.

## List of abbreviations

LRRH: Lira Regional Referral Hospital
LUREC: Lira University Research Ethics Committee
MOH: Ministry of Health
PSCS: Premarital Sickle Cell Screening
SCD: Sickle Cell Disease
SCT: Sickle Cell Trait
UNCST: Uganda National Council for Science and Technology
WHO: World Health Organization
SDG: Sustainable Developmental Goals

## Declarations

### Ethics approval and consent to participate

This study received ethical approval from the Lira University Research and Ethics Committee (LUREC) under reference number LUREC-2024-291, and administrative clearance was obtained from Lira Regional Referral Hospital (LRRH). The research adhered strictly to ethical standards regarding human subjects. Written informed consent was obtained from all participants.

### Consent for publication

Not applicable. This manuscript does not contain any individual person’s data, images, or videos.

### Availability of data and materials

The datasets analysed during the current study are not publicly available due to participant confidentiality but are available from the corresponding author upon reasonable request.

### Authors’ contributions

AN conceptualized the study, developed the research protocol, collected and analyzed the data, interpreted the results, and drafted the manuscript. JWB (Jairus Wangusa Byawele) guided in data analysis and manuscript development. BA (Bridget Akiror) and EO (Emmanuel Ogwal) provided guidance and support whenever needed during the research process. MSO (Dr. Marc Sam Opollo) and MA (Maxson Anyolitho) offered supervision, expert guidance, and critical revisions throughout the development of the study and the writing process. All authors read and approved the final version of the manuscript.

## Data Availability

The datasets analyzed during the current study are not publicly available due to participant confidentiality but are available from the corresponding author upon reasonable request.

## Acknowledgements

The author wishes to acknowledge the Almighty God for the strength, good health, and wisdom granted throughout the research journey. Sincere gratitude is extended to the academic supervisor, Dr. Marc Sam Opollo and Dr. Maxson Anyolitho for his invaluable mentorship, and to Jairus Wangusa Byawele, Bridget Akiror, and Emmanuel Ogwal for their constructive feedback and unwavering support. Special appreciation goes to the Faculty of Public Health at Lira University for providing the academic and institutional foundation for this research.

The author also acknowledges the administration and staff of Lira Regional Referral Hospital, particularly those in the pediatric and sickle cell departments, for their cooperation and support during data collection. Lastly, heartfelt thanks go to family and friends for their constant encouragement and moral support throughout the study period.

